# Economic cost of paediatric heart disease in the United Kingdom: A national tertiary healthcare and societal cost-of-illness analysis using real-world data

**DOI:** 10.64898/2026.09.29.26362732

**Authors:** Rebecca Naylor, Anna Buylova, Siddesh Shetty, Thomas Witter, Samara Grant, Nick Byrne, Kuberan Pushparajah

**Affiliations:** Health Innovation Network South London, Guy’s and St Thomas’s Hospital, London, UK; Department of Paediatric Cardiology, Evelina Children’s Hospital, London, UK; NIHR HealthTech Research Centre Cardiovascular and Respiratory medicine: Heart disease in children, Guy’s and St Thomas’ NHS Foundation Trust and King’s College London, London, UK; Medical Physics & Clinical Engineering, Guy’s and St Thomas’ NHS Foundation Trust, London, UK; School of Biomedical Engineering & Imaging Sciences, King’s College London, London, UK

**Keywords:** Congenital heart disease, cost-of-illness, paediatric cardiology, economic cost

## Abstract

**Background/Aims:** This work aimed to estimate the annual tertiary healthcare and societal costs associated with paediatric congenital heart disease (CHD) in the United Kingdom using real-world data from a Level 1 cardiac surgical centre in the National Health Service (NHS).

**Methods:** A prevalence-based cost-of-illness analysis was conducted using patient-level inpatient and outpatient activity data from Evelina London Children’s Hospital between 1 June 2023 and 31 May 2024. Costs were estimated from NHS and societal perspectives and extrapolated to national level using National Congenital Heart Disease Audit (NCHDA) procedural activity data. Healthcare costs included tertiary centre care costs. Societal costs included parental productivity losses and impacts on lifetime earnings for children. Deterministic sensitivity analysis explored parameter uncertainty.

**Results:** Approximately 9.6% of UK paediatric CHD procedures were undertaken at Evelina London Children’s Hospital at a cost of £15.0 million per annum, corresponding to a national estimate of £156.4 million. Single-ventricle procedures accounted for 4.8% of inpatient activity but 19.4% of inpatient costs. Sensitivity analysis produced national healthcare cost estimates ranging from £125.1 million to £187.7 million.

National productivity losses associated with parental absence from work were estimated at £82.4 million annually. Individuals with CHD were estimated to experience a mean reduction in lifetime earnings of £81,509.

**Conclusion:** Paediatric CHD imposes a substantial economic cost on the UK healthcare system and wider society. This study provides the first real-world national estimate of the economic impact of paediatric CHD in the UK and may inform future service planning, research investment, and innovation adoption strategies.

**Key Learning points:** *1. What is already known*

• Congenital heart disease (CHD) is the most common congenital abnormality worldwide and requires substantial healthcare utilisation throughout childhood, including outpatient monitoring, hospital admissions, surgical and catheter interventions.

• Existing evidence suggests that CHD is associated with high healthcare costs and wider societal impacts; however, available economic burden studies have been conducted in the United States and are not generalisable to the UK healthcare system.

• The broader economic consequences of paediatric CHD, including parental productivity losses and impacts on future employment and earnings, remain poorly quantified in the UK.

*2. What this study adds*

• This study provides, to our knowledge, the first UK-wide estimate of the economic burden of paediatric CHD using real-world patient-level activity and cost data from a large volume tertiary centre.

• Annual direct NHS costs associated with paediatric CHD care at tertiary settings were estimated at £156.4 million nationally, with an additional £82.4 million in parental productivity losses, demonstrating a substantial societal burden beyond healthcare expenditure alone.

• Infants and patients with severe CHD requiring single-ventricle procedures accounted for a disproportionately large share of costs, highlighting important targets for future service improvement, research investment, and innovation.

## Introduction

Congenital heart disease (CHD) is the most common congenital abnormality worldwide, affecting approximately nine children per 1,000 live births (1). CHD encompasses a heterogeneous group of structural cardiac abnormalities ranging from relatively minor defects requiring limited intervention to complex lesions necessitating multiple surgical procedures and lifelong specialist care. Advances in diagnosis, surgery, intensive care, and long-term management have substantially improved survival over recent decades, resulting in a growing population of children and adolescents living with CHD and requiring ongoing healthcare support (2).

The burden of paediatric CHD extends beyond its clinical consequences. Children with CHD require primary, secondary and tertiary care, with increasing levels of specialisation. In the UK, this is delivered within Congenital heart disease networks of care, which consist of Specialist Children’s Surgical Centres (Level 1), Children’s Cardiology Centres (Level 2) and Local Children’s Cardiac Centres (Level 3) (3).

Patients can enter the paediatric cardiac pathway through a prenatal diagnosis, physical examination of an asymptomatic or symptomatic infant, or referrals from the obstetric department, neonatal centre or primary care (3). The care pathway can include repeated outpatient surveillance, diagnostic investigations, hospital admissions, medical management, surgical and catheter interventions, and multidisciplinary follow-up throughout childhood and adolescence. Complex forms of CHD are associated with prolonged hospitalisation, higher rates of complications, and greater healthcare resource utilisation than less severe disease (4).

Consequently, CHD represents a substantial economic burden for healthcare systems as well as for affected children and their families.

In addition to direct healthcare costs, paediatric CHD may result in significant societal costs. Parents and caregivers often provide substantial informal care and may require time away from employment to attend clinic appointments, support hospital admissions, and assist with postoperative recovery (5). Furthermore, health- related disruptions during childhood can influence educational attainment and long- term employment opportunities, potentially affecting lifetime earnings among individuals living with CHD (6). These broader economic consequences are increasingly recognised as important components of disease burden but remain poorly quantified within the United Kingdom (UK).

Despite increasing policy focus on improving outcomes for children and adolescents, paediatric healthcare has historically received relatively limited research and commercial investment compared with adult healthcare (7). This challenge is evident within paediatric cardiology, where the relatively small patient population may reduce incentives for innovation and investment (8). Robust evidence describing the economic burden of paediatric CHD is therefore important to support healthcare planning, prioritisation of research funding, and the evaluation of emerging technologies and models of care.

Existing evidence describing the economic burden of CHD is largely derived from studies conducted in the United States, with limited data available that are specific to the UK healthcare system (9, 10). Differences in healthcare delivery models, reimbursement mechanisms, and social care structures limit the generalisability of international estimates to the UK setting. Furthermore, previous studies have frequently focused on specific procedures, disease subtypes, or healthcare costs alone, without capturing broader societal impacts. As a result, there remains a significant evidence gap regarding the overall economic burden of paediatric CHD in the UK.

Guy’s and St Thomas’ NHS Foundation Trust (GSTT), through Evelina London Children’s Hospital and the Royal Brompton Hospital, is the largest provider of paediatric CHD care in the UK (11). Using real-world patient-level activity and cost data from Evelina London Children’s Hospital, this study aimed to estimate the annual economic burden of paediatric CHD in the UK from both a tertiary healthcare centre system and societal perspective. By providing national estimates of economic burden, this study seeks to inform future research, policy development, service planning, and investment in innovative approaches to paediatric CHD care.

## Methods

### Study design

A cost-of-illness study was undertaken to estimate the annual economic burden of paediatric CHD in the UK. This included congenital, inherited and acquired heart diseases in patients up to and including 18 years of age as per National Congenital Heart Disease Audit (NCHDA) definition of CHD (11). Methodology followed that of Schnitzler et al, guidelines for cost-of-illness studies (12). The analysis used a bottom-up costing approach and adopted both a healthcare system perspective and a broader societal perspective and applied a one-year time horizon. The study was conducted in two stages. Stage one estimated the direct healthcare costs associated with paediatric CHD from the National Health Service (NHS) perspective, while stage two expanded the analysis to include wider societal costs arising from parental productivity losses and the impact of CHD on lifetime earnings for children and adolescents. All analysis was undertaken using Microsoft Excel 365. Results are reported as annual costs at both the provider-level and the estimated national level, stratified by age specific groups. As the time horizon of the analysis is one year, discounting of costs was not applied (13).

### Data sources and study population

Individual patient-level inpatient and outpatient activity data were obtained from Evelina London Children’s Hospital, GSTT. Data were supplied by the Trust’s Business Intelligence team and included healthcare activity, appointment type, attendance information, and associated costs. The analysis used activity occurring between 1 June 2023 and 31 May 2024, representing the most recent period for which consistent inpatient and outpatient datasets were available. Local institutional approval for the use of data was acquired as part of a locally registered quality improvement project 17575. The study population included children and adolescents aged under 19 years receiving care for CHD. Key data inputs used in the analysis are summarised in Table 1.

**Table 1:**
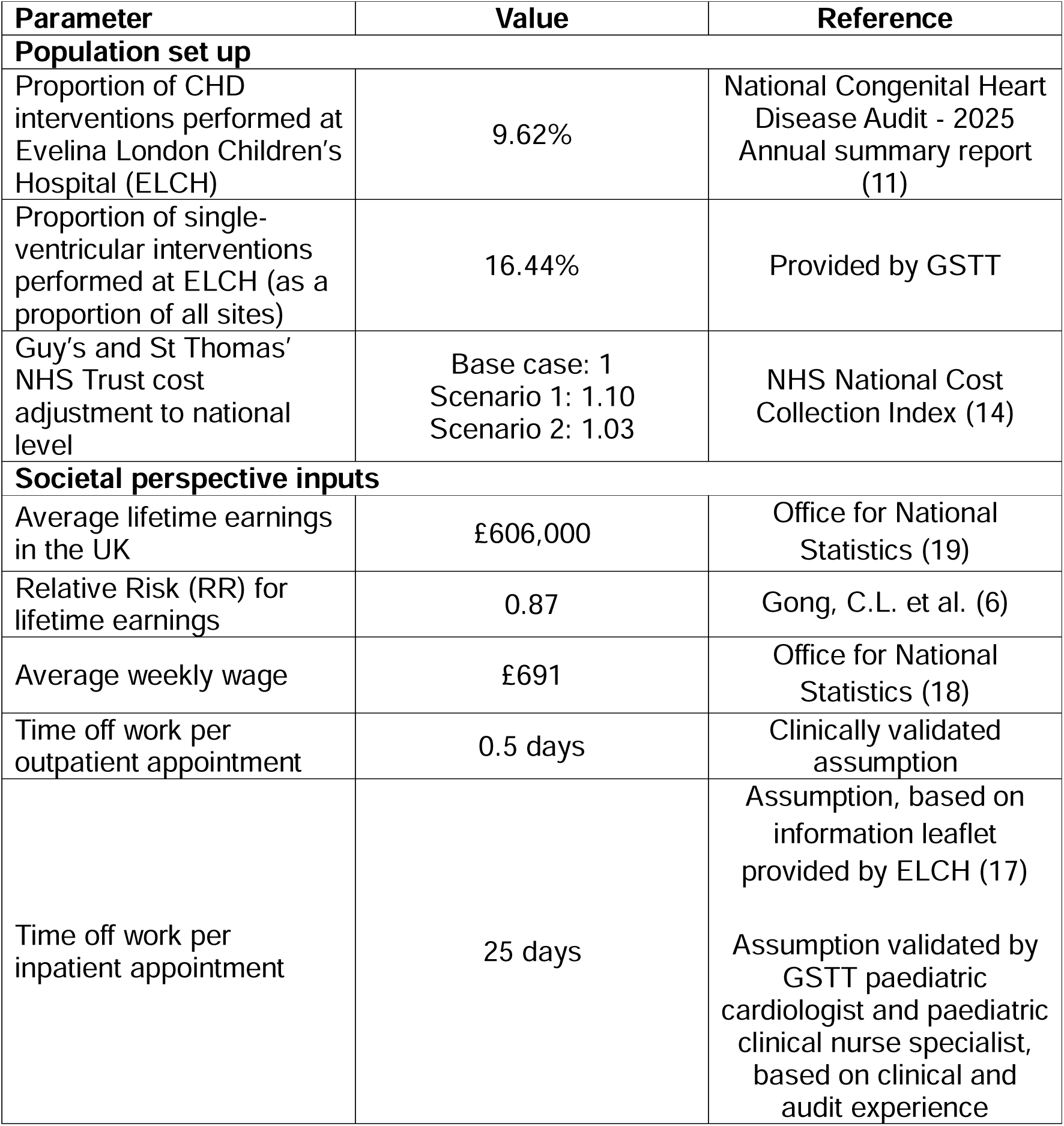
Model inputs.

### NHS perspective analysis

Healthcare resource use included both inpatient admissions and outpatient attendances. Outpatient costs were based on contract monitoring prices (cost charged for services), while inpatient costs were based on activity-related income received by the Trust.

National estimates were generated by extrapolating Evelina London Children’s Hospital activity using procedural data from the NCHDA (11). A UK perspective was adopted to align with the geographic reporting framework used by the NCHDA and to facilitate comparison with national congenital cardiac activity statistics. The Republic of Ireland was excluded from the analysis; therefore, estimates relate to patients receiving care within English congenital cardiac centres included in the NCHDA. According to 2023/24 NCHDA data, Evelina London Children’s Hospital accounted for 9.6% of paediatric CHD procedures performed nationally during 2023/24. A multiplier of 10.4 was therefore used to estimate NHS costs at the UK level. For the single-ventricle subgroup, a separate, specific multiplier based on national procedural activity (16.4%, provided by GSTT), was applied.

To assess uncertainty surrounding cost extrapolation, two scenario analyses were conducted using the NHS National Cost Collection Index (NCCI) (14). Scenario 1 adjusted costs using the GSTT-specific NCCI, while Scenario 2 applied a weighted average NCCI derived from UK specialist paediatric cardiac centres. These analyses explored the impact of variation in provider-level cost structures on national cost estimates.

### Societal perspective analysis

The societal perspective incorporated productivity losses associated with parental caregiving and the longer-term impact of CHD on patient lifetime earnings. A human capital approach was adopted to estimate productivity losses (15), as is traditional in cost-of-illness methods. Informal care costs were estimated applying opportunity cost approach (16).

For outpatient activity, productivity losses were estimated using a clinically guided assumption of 0.5 days of work absence per appointment. For inpatient activity, parental productivity losses combined hospital length of stay with an additional five- week recovery period following discharge, based on clinical expert opinion and patient information provided by Evelina London Children’s Hospital (17). Productivity losses were valued using average weekly earnings reported by the Office for National Statistics (ONS) (18). Average weekly income from May 2024 was used, in line with the analysis period.

The impact of CHD on lifetime earnings was estimated using a previously published microsimulation study from the United States, which reported lower lifetime earnings among individuals living with congenital heart disease (6). The relative reduction in earnings identified in that study was applied to UK estimates of lifetime earnings to generate a UK-specific estimate of long-term economic impact. This estimate was taken from ONS for the year 2022, which was the most recent estimate at the time of analysis (19).

### Subgroup analysis

Resource use and costs were analysed across five age groups: <1 year, 1–4 years, 5–9 years, 10–14 years, and 15–18 years. A predefined subgroup analysis was undertaken for patients undergoing single-ventricle procedures. This group was selected because single-ventricle physiology is associated with particularly complex management pathways and substantial healthcare resource utilisation. Healthcare costs and societal costs were estimated separately for this subgroup using procedure-specific national extrapolation factors and age-specific length-of-stay data from the Evelina London Children’s Hospital dataset.

### Sensitivity analysis

Deterministic sensitivity analysis (DSA) was performed to evaluate the robustness of model results to uncertainty in key parameters. Consistent with standard practice in economic evaluations, the number of appointments and attendances was varied by ±20%. The impact of these changes on estimates of healthcare costs, societal costs, and lifetime earnings losses was examined to identify key drivers of the overall economic burden.

### Quality assurance

All datasets and summary outputs were independently validated using RStudio (Version 2025.09.1+401). Descriptive statistics were used to summarise healthcare utilisation and associated costs. The methodological approach and analysis conform to the Consolidated Health Economic Evaluation Reporting Standards (CHEERS) 2022 checklist (20).

## Results

### Study population and healthcare activity

During the study period (1 June 2023 to 31 May 2024), 8,464 children and adolescents with CHD received care at Evelina London Children’s Hospital through both outpatient and inpatient services. The cohort included patients across all paediatric age groups, with the greatest healthcare utilisation observed among infants and younger children. A total of 1,935 inpatient admissions and 9,400 outpatient appointments were identified between 8,464 patients, representing a substantial volume of specialist CHD care delivered over the study period. Outpatient activity accounted for the majority of healthcare interactions, whereas inpatient admissions accounted for the greatest proportion of healthcare expenditure. Infants aged <1 year exhibited the highest resource utilisation and cost burden, reflecting the intensive treatment requirements associated with complex congenital heart lesions in the first year of life. Table 2 summarises patient characteristics and healthcare activity by age group.

**Table 2:** Annual appointment data (1 June 2023-31 May 2024)

| Parameter | Base-case |  | Single<br>ventricle |
| --- | --- | --- | --- |
| Age | Inpatient | Outpatient | Inpatient |
| **Number of people in cohort** |  |  |  |
| Age < 1 years | 590 | 897 | 53 |
| Age 1 ≤ 5 years | 342 | 1599 | 12 |
| Age 5 to ≤ 10 years | 241 | 1698 | 5 |
| Age 10 to ≤ 15 years | 270 | 2047 | 4 |
| Age 15 to ≤ 18 years | 128 | 1289 | 2 |
| **Total number of appointments per age group** |  |  |  |
| Age < 1 years | 794 | 1411 | 54 |
| Age 1 ≤ 5 years | 422 | 1898 | 12 |
| Age 5 to ≤ 10 years | 264 | 1927 | 5 |
| Age 10 to ≤ 15 years | 311 | 2506 | 4 |
| Age 15 to ≤ 18 years | 144 | 1658 | 2 |

### Healthcare costs

Total annual healthcare costs incurred by the Evelina London Children’s Hospital study cohort were estimated at £15.0 million. Of this, more than £7.0 million of the cost was associated with the care of those aged less than 1 year old. Extrapolation using NCHDA procedural activity data indicated that Evelina London Children’s Hospital accounted for approximately 9.6% of national paediatric CHD procedures, resulting in an estimated annual UK healthcare burden of £156.4 million.

Inpatient admissions represented the largest component of healthcare expenditure. Average costs varied considerably by age group, with infants (<1 year) accounting for a disproportionate share of total expenditure (51.1% of cost borne by 37.6% of cohort) due to higher rates of surgical intervention and prolonged hospitalisation.

National annual healthcare costs by age group are presented in Table 3.

**Table 3:** Annual healthcare costs by age group (GBP, 1 June 2023-31 May 2024)

| Type of cost | Value |  |
| --- | --- | --- |
| **Healthcare costs – per person** | **Inpatient** | **Outpatient** |
| Age < 1 years | £11,499 | £313 |
| Age 1 ≤ 5 years | £7,699 | £218 |
| Age 5 to ≤ 10 years | £6,528 | £208 |
| Age 10 to ≤ 15 years | £5,785 | £229 |
| Age 15 to ≤ 18 years | £5,761 | £237 |
| **Healthcare costs – per episode** | **Inpatient** | **Outpatient** |
| Age < 1 years | £8,544 | £199 |
| Age 1 ≤ 5 years | £6,240 | £183 |
| Age 5 to ≤ 10 years | £5,959 | £183 |
| Age 10 to ≤ 15 years | £5,022 | £187 |
| Age 15 to ≤ 18 years | £5,121 | £184 |
| **Healthcare costs – Evelina cohort** | **Inpatient** | **Outpatient** |
| Age < 1 years | £6,784,309 | £280,890 |
| Age 1 ≤ 5 years | £2,633,133 | £347,995 |
| Age 5 to ≤ 10 years | £1,573,202 | £353,490 |
| Age 10 to ≤ 15 years | £1,561,887 | £468,751 |
| Age 15 to ≤ 18 years | £737,357 | £305,527 |
| **Total** | £13,289,888 | £1,756,654 |
|  | **£15,046,542** |  |
| **Healthcare costs – National level estimate** | **Inpatient** | **Outpatient** |
| Age < 1 years | £70,510,511 | £2,919,343 |
| Age 1 ≤ 5 years | £27,366,611 | £3,616,777 |
| Age 5 to ≤ 10 years | £16,350,564 | £3,673,887 |
| Age 10 to ≤ 15 years | £16,232,969 | £4,871,816 |
| Age 15 to ≤ 18 years | £7,663,478 | £3,175,391 |
| **Total** | £138,124,134 | £18,257,214 |
|  | **£156,381,347** |  |

### Societal costs

#### Parental Productivity Losses

Paediatric CHD generated considerable indirect costs through parental and caregiver productivity losses. Based on assumptions regarding time required for hospital attendance and recovery support following hospitalisation, annual productivity losses attributable to paediatric CHD were estimated at £82.4 million nationally.

The majority of productivity losses arose from inpatient admissions (91.8%), reflecting both prolonged hospital stays and additional caregiver responsibilities during recovery periods. Productivity losses associated with outpatient appointments represented a smaller but meaningful component of overall societal costs of £6.7 million per annum. Results of annual societal cost analyses are presented in Table 4.

**Table 4:** Annual societal costs by age group.

| Type of cost | Value |  |
| --- | --- | --- |
|  | Inpatient | Outpatient |
| **Societal costs – Evelina cohort** |  |  |
| Age < 1 years | £3,138,798 | £97,500 |
| Age 1 ≤ 5 years | £1,535,331 | £131,152 |
| Age 5 to ≤ 10 years | £954,133 | £133,156 |
| Age 10 to ≤ 15 years | £1,125,501 | £173,165 |
| Age 15 to ≤ 18 years | £523,502 | £114,568 |
| **Total** | £7,277,264 | £649,540 |
|  | **£7,926,804** |  |
| **Societal costs – National level estimate** |  |  |
| Age < 1 years | £32,622,081 | £1,013,336 |
| Age 1 ≤ 5 years | £15,956,961 | £1,363,084 |
| Age 5 to ≤ 10 years | £9,916,469 | £1,383,911 |
| Age 10 to ≤ 15 years | £11,697,527 | £1,799,730 |
| Age 15 to ≤ 18 years | £5,440,844 | £1,190,723 |
| **Total** | £75,633,882 | £6,750,783 |
|  | **£82,384,665** |  |

#### Lifetime Earnings Impact

The estimated mean reduction in lifetime earnings was £81,509 per individual with CHD compared with the general population. When considered across the national CHD population, these losses represent a substantial additional societal burden extending beyond childhood healthcare utilisation.

### Scenario analysis and single ventricle group

Application of alternative cost extrapolation methods demonstrated that national cost estimates were sensitive to assumptions regarding provider-specific cost structures. Using the GSTT National Cost Collection Index (NCCI), total annual national healthcare costs were estimated at £142.3 million. Applying a weighted average NCCI derived from specialist paediatric cardiac centres generated a higher estimate of £152.0 million. Despite variation across scenarios, all analyses consistently demonstrated a substantial national economic burden associated with paediatric CHD.

Patients undergoing single-ventricle procedures represented a relatively small proportion of overall activity but generated a disproportionate share of healthcare costs. Single-ventricle procedures accounted for approximately 4.8% of inpatient activity in the Evelina cohort, but was estimated to account for approximately 19.4% of total inpatient expenditure. This reflects the complexity of care pathways associated with single-ventricle physiology, including extended hospital stays, multiple surgical procedures, and increased postoperative resource requirements. National extrapolation suggested that severe CHD requiring single-ventricle intervention accounted for a substantial proportion of the overall economic burden despite representing a minority of patients. Healthcare and societal costs for the single-ventricle subgroup are summarised in Table 5.

**Table 5:** Sub-group (single ventricle) analysis results.

| Type of cost | Value |  |
| --- | --- | --- |
| Healthcare costs – per person | Per person | Per episode |
| Age < 1 years | £36,289 | £35,617 |
| Age 1 ≤ 5 years | £28,688 | £28,688 |
| Age 5 to ≤ 10 years | £28,237 | £28,237 |
| Age 10 to ≤ 15 years | £30,084 | £30,084 |
| Age 15 to ≤ 18 years | £26,271 | £26,271 |
| Healthcare costs – Evelina cohort | Inpatient cost | Proportion (%) |
| Age < 1 years | £1,923,337 | 28.35% |
| Age 1 ≤ 5 years | £344,254 | 13.07% |
| Age 5 to ≤ 10 years | £141,185 | 8.97% |
| Age 10 to ≤ 15 years | £120,336 | 7.70% |
| Age 15 to ≤ 18 years | £52,541 | 7.13% |
| **Total** | **£2,581,653** | **19.43%** |
| Healthcare costs – National level estimate | Inpatient cost |  |
| Age < 1 years | £11,700,300 |  |
| Age 1 ≤ 5 years | £2,094,212 |  |
| Age 5 to ≤ 10 years | £858,875 |  |
| Age 10 to ≤ 15 years | £732,044 |  |
| Age 15 to ≤ 18 years | £319,624 |  |
| **Total** | **£15,705,056** |  |
| Societal costs – Evelina cohort | Societal cost |  |
| Age < 1 years | £304,785 |  |
| Age 1 ≤ 5 years | £50,167 |  |
| Age 5 to ≤ 10 years | £20,177 |  |
| Age 10 to ≤ 15 years | £18,242 |  |
| Age 15 to ≤ 18 years | £7,463 |  |
| **Total** | **£400,834** |  |
| Societal costs – National level estimate | Societal cost |  |
| Age < 1 years | £1,854,107 |  |
| Age 1 ≤ 5 years | £305,180 |  |
| Age 5 to ≤ 10 years | £122,745 |  |
| Age 10 to ≤ 15 years | £110,975 |  |
| Age 15 to ≤ 18 years | £45,399 |  |
| **Total** | **£2,438,405** |  |

### Sensitivity analysis

Deterministic sensitivity analysis demonstrated that estimates were most sensitive to assumptions regarding volumes of healthcare utilisation, particularly inpatient admissions and outpatient attendance rates. Variation of key utilisation parameters by ±20% produced national healthcare cost estimates ranging from £125.1 million to £187.7 million annually. Despite this uncertainty, paediatric CHD remained associated with a substantial economic burden under all tested assumptions. Parental productivity losses and lifetime earnings estimates showed similar patterns, with results remaining consistently high across all modelled scenarios. The results of this analysis are presented in Table 6.

**Table 6:** Deterministic sensitivity analysis results.

| **Type of cost** | **Base-case** | **Upper value** | **Lower value** |
| --- | --- | --- | --- |
| Healthcare costs – Evelina cohort | £15,046,542 | £18,055,851 | £12,037,234 |
| Healthcare costs – National level | £156,381,347 | £187,657,617 | £125,105,078 |
| Societal costs – Evelina cohort | £7,926,804 | £9,512,165 | £6,341,443 |
| Societal costs – National level | £82,384,665 | £98,861,598 | £65,907,732 |
| Lifetime earnings loss per person | £81,509 | £97,811 | £65,207 |

## Discussion

This study provides, to our knowledge, the first UK-wide estimate of the economic burden of paediatric congenital heart disease (CHD) using real-world hospital activity and costing data. The analysis demonstrated that paediatric CHD constitutes a substantial cost to both the healthcare system and wider society. Annual direct healthcare costs were estimated at £156.4 million nationally, while parental productivity losses associated with healthcare attendance and recovery periods were estimated at £82.4 million annually. In addition, children and adolescents with CHD were estimated to experience a mean reduction in lifetime earnings of £81,509 (13.0%) compared with the general population.

A key finding was the concentration of costs within inpatient care. Although outpatient activity represented the majority of healthcare contacts, inpatient care accounted for the majority of total healthcare expenditure. This reflects the resource- intensive nature of paediatric CHD care, particularly among infants who frequently require diagnostic investigations, surgical and transcatheter interventions, intensive care support, and prolonged hospitalisation during the first year of life. This may also be reflective of the data sample, where primary care costs were excluded. The subgroup analysis further demonstrated that severe disease disproportionately contributes to healthcare expenditure. Patients undergoing single-ventricle procedures represented less than 5% of inpatient activity but accounted for almost one-fifth of total inpatient costs. This is in line with existing research conducted in the UK, where 5.9% of those who received an intervention for CHD between 2000 and 2018 had a single-ventricle defect (21). These findings highlight the substantial resource requirements associated with complex CHD and suggest that interventions targeting patients with the greatest clinical complexity may offer particularly large opportunities for improving efficiency and outcomes. It is also recognised that these costs are highest in the earliest year of their treatment pathway (22).

### Comparison with existing literature

There is currently limited evidence quantifying the economic burden of paediatric CHD within the UK, making direct comparisons challenging. Most published studies have originated from the United States and have focused on specific disease subgroups, surgical procedures, or healthcare settings rather than the broader paediatric CHD population (23, 24). Studies from the United States have consistently identified CHD as one of the costliest childhood conditions due to high rates of hospitalisation and specialist care utilisation (23). The findings of the present study are consistent with this literature and confirm that CHD represents a major source of healthcare expenditure within paediatric cardiology.

To contextualise the findings, the estimated annual NHS cost of paediatric CHD (£156.4 million) lies between published estimates for other childhood conditions. Recent evidence suggests an annual healthcare cost of approximately £300 million for paediatric obesity and approximately £60 million for paediatric asthma in the UK (25, 26). The present findings therefore indicate that paediatric CHD represents one of the most economically significant chronic childhood conditions despite its relatively low prevalence. The disproportionate burden associated with severe disease observed in the current analysis is also consistent with previous studies of paediatric cardiac surgery. Hudson et al. reported substantially higher hospital costs among patients experiencing postoperative morbidity following paediatric cardiac surgery, highlighting the importance of disease complexity and complications as key drivers of expenditure (24). Our findings extend this evidence by demonstrating the broader national implications of resource utilisation across the paediatric CHD population.

### Strengths and limitations

This study has several strengths. Firstly, the analysis provides to our knowledge the first UK-wide estimate of the economic burden of paediatric CHD, using real-world patient-level activity and costing data obtained from a large provider of paediatric CHD care in the UK. The use of routinely collected healthcare data enhances the relevance and generalisability of findings to contemporary clinical practice. Secondly, the analysis adopted both healthcare and societal perspectives, capturing not only tertiary healthcare expenditure but also parental productivity losses and the longer- term impact of CHD on lifetime earnings. This broader perspective allowed for a more comprehensive assessment of economic burden than studies restricted to direct healthcare costs alone. Thirdly, national extrapolation was based on procedural activity reported through the NCHDA, providing a transparent and reproducible methodology for estimating UK-wide costs. Finally, the study incorporated subgroup and sensitivity analyses, which demonstrated the disproportionate economic burden associated with single-ventricle CHD and confirmed the robustness of findings across a range of assumptions.

Several limitations should be considered when interpreting the results. National estimates were derived from data collected at a single provider and extrapolated using procedural activity data. Although Evelina London Children’s Hospital is a designated Level 1 cardiac surgical centre in the UK, patient case mix and resource use may differ across centres. In addition, the analysis was restricted to tertiary care and did not include activity delivered in primary care, Level 2 and Level 3 cardiac services, community care, social care and education services. Although the impact to lifetime earnings was estimated, evidence shows that children with CHD have greater special educational need and require additional support (28). Additionally, transplant related costs were not included as these are delivered by specially commissioned national services. The analysis adopted a UK perspective in line with the NCHDA. However, Scotland has not contributed data to the NCHDA since 2021 (11). As a result, activity undertaken within Scottish congenital cardiac services is not fully captured within the national extrapolation. Care delivered at congenital cardiac centres outside of England and Wales may therefore not be captured. Consequently, the estimates are likely an underestimate of the true economic burden of paediatric CHD. The societal analysis relied on assumptions regarding parental productivity losses and used US evidence to inform lifetime earnings impacts due to the absence of UK-specific data. Finally, the analysis did not capture health-related quality of life or psychological impacts on patients and caregivers, which represent important dimensions of burden beyond economic costs.

### Societal and policy implications

While economic evaluations often focus on healthcare expenditure alone, caring for children with CHD frequently requires substantial parental involvement. Hospital admissions, recovery periods, and routine outpatient follow-up can result in repeated absences from employment and consequent productivity losses. The estimated annual productivity loss of £82.4 million highlights the wider economic consequences of paediatric CHD that are not directly captured within healthcare budgets.

Published evidence regarding the long-term economic consequences of congenital heart disease suggests that individuals living with CHD may experience substantial reductions in earning potential over their lifetime (23). The estimated reduction in lifetime earnings further emphasises the long-term impact of CHD beyond childhood. Although advances in treatment have dramatically improved survival, individuals with CHD may still experience educational disruption, reduced labour market participation, and barriers to employment. These findings suggest that improving outcomes in paediatric CHD has the potential to generate broader societal benefits extending beyond reductions in healthcare expenditure.

These results support the case for greater investment in paediatric health research and innovation. Children and adolescents have historically received a disproportionately small share of healthcare research and innovation funding compared with adults, despite accounting for a substantial proportion of healthcare need (8). By quantifying the economic burden associated with paediatric CHD, this study provides evidence that investments aimed at improving outcomes, reducing complications, and streamlining care pathways may generate substantial returns for both the healthcare system and wider economy. Such evidence may be particularly valuable for supporting grant applications, business cases, and commercial investment in paediatric cardiovascular technologies. Given the concentration of costs in infancy and among patients with severe disease, interventions targeted towards these groups may have the greatest potential economic impact.

The study highlights the need for improved national data collection and reporting. Current estimates were derived from a single specialist provider and extrapolated nationally because comprehensive UK-wide resource-use data are not routinely available. The development of linked national datasets incorporating hospital care, primary care, community services, educational outcomes, and socioeconomic indicators would improve understanding of the full burden of CHD and support more evidence-based policy decisions. In particular, improved collection of ethnicity, deprivation, and geographic access data would enable evaluation of health inequalities and support more equitable service delivery.

### Future research

Future research should seek to validate these findings using data from multiple congenital cardiac centres across the UK. Additional work is also needed to quantify costs incurred within primary care, community services, education, and social care.

The development of UK-specific evidence regarding educational outcomes, employment participation, and long-term productivity impacts among individuals with CHD would strengthen future societal analyses. Research exploring health inequalities, including the role of ethnicity, deprivation, and geographical access to specialist services, may help identify opportunities to improve outcomes and reduce disparities in care.

Finally, future studies should incorporate patient-reported outcomes and quality-of- life measures to provide a more comprehensive assessment of the burden of paediatric CHD and the potential value of interventions designed to improve care.

## Conclusion

Paediatric congenital heart disease is associated with considerable healthcare and societal costs in the UK. National annual healthcare costs exceeded £156 million, with further costs arising through parental productivity losses and reduced lifetime earnings. These findings provide an important foundation for future economic evaluations and innovation strategies aimed at improving outcomes and reducing the burden of paediatric CHD.

## Funding

This paper represents independent research supported by the National Institute for Health and Care Research (NIHR) HealthTech Research Centre (HRC) in Cardiovascular and Respiratory Medicine at Guy’s and St Thomas’ NHS Foundation Trust and King’s College London. The views expressed are those of the author(s) and not necessarily those of the NHS, the NIHR or the Department of Health and Social Care.

## Data Availability

The research data supporting this publication are stored securely by Guy's and St Thomas' NHS Foundation Trust. Restrictions apply to the availability of these data, which were used under license for the current study, and so are not publicly available.

## Notes

### Competing Interest Statement

The authors have declared no competing interest.

### Author Declarations

The clinical audit process within Guy's and St Thomas' NHS Foundation Trust gave approval for this work with audit number 17575. The datasets used in the study were pseudonymised by the BI team at the trust before we received them.

